# Artificial Intelligence Enabled Phenogrouping of Heart Failure with Preserved Ejection Fraction Depicts Early and End-Stage Trajectories

**DOI:** 10.1101/2025.06.09.25329306

**Authors:** Samuel Brown, Fardad Soltani, Jack Wu, Matthew Ryan, Brett S Bernstein, Brian Tam To, Tom Searle, Maleeha Rizvi, Natalie Fairhurst, George Kaye, Ranu Baral, Dhanushan Vijayakumar, Daksh Mehta, Zeeshan Khawaja, Phil Chowienczyk, James Teo, Richard JB Dobson, Daniel I Bromage, Gerald Carr-White, Thomas F Lüscher, Ali Vazir, Theresa A McDonagh, Jessica Webb, Ajay M Shah, Christopher A Miller, Dhruva Biswas, Kevin O’Gallagher

**Author notes:** **Corresponding author:** Dr Kevin O’Gallagher, PhD, The James Black Centre, School of Cardiovascular and Metabolic Medicine & Sciences, British Heart Foundation Centre of Research Excellence, King’s College London, 125 Coldharbour Lane, London, UK, SE5 9NU. Joint senior authors (Dhruva Biswas, Kevin O’Gallagher).

## Abstract

**Background:** Heart failure with preserved ejection fraction is challenging to diagnose, precluding the initiation of prognostic medications. A deeper understanding of HFpEF phenotypes and trajectories could address this, however previous studies analyzed trial cohorts, limiting generalizability. Here we apply artificial intelligence (AI) algorithms to longitudinal electronic health record data (EHR) to characterize reproducible real-world HFpEF phenogroups.

**Methods:** We applied natural language processing (NLP) to extract structured and unstructured data from EHRs at a multi-site hospital network, including comorbidities, echocardiographic measurements, and all-cause mortality outcomes. HFpEF cases were identified by applying contemporaneous guideline-directed criteria. Phenogroups were derived at the first documented mention of ‘heart failure’ using latent class analysis, then reassigned annually to quantify the frequency and directionality of transitions. External validation was performed at an independent hospital network.

**Results:** We identified 2,223 patients (median age 75 years; 60% female) meeting HFpEF diagnostic criteria, with a median follow-up of 4 years. A clinician-recorded HFpEF diagnosis was present for only 10.4%. Four phenogroups were identified: Elderly-Atrial Dysfunction (32%), Cardio-Renal-Metabolic (23%), Obesity-Predominant (24%), and Young-Low Comorbidity (22%). The Young-Low Comorbidity group had the lowest rate of clinician-assigned HFpEF diagnosis (3%), and more than half of patients transitioned phenogroups within five years, suggesting this may represent an early-stage HFpEF phenotype. By contrast, Elderly-Atrial Dysfunction and Cardio-Renal-Metabolic groups displayed stable group membership (<10% transitioned), with high mortality rates (adjusted hazard ratio = 1.30 [1.02–1.67] and 1.49 [1.18–1.87] respectively). Phenogroup reproducibility was validated externally (n=3,349), demonstrating consistent clinical features and survival trends.

**Conclusion:** AI-enabled phenotyping of real-world data identified reproducible, prognostically significant HFpEF phenogroups. The identification of an underrecognized, early-stage subgroup underscores a critical opportunity for earlier diagnosis and targeted intervention to alter the clinical trajectory.

**Clinical perspectives:** *What is new?:* Heart failure with preserved ejection fraction (HFpEF), as a heterogenous syndrome, can be characterized into four reproducible phenogroups, including an early-HFpEF phenotype.

*What are the clinical implications?:* Future research could build on implementation of scalable, EHR-enabled diagnostic strategies for early HFpEF detection. Longitudinal studies examining the impact of intervention on early phenogroups may provide valuable insights into modifying disease progression.

## Introduction

Heart failure with preserved ejection fraction (HFpEF) represents a critical public health challenge, accounting for approximately half of all heart failure cases.^1^ Yet, we and others have shown that HFpEF remains significantly underrecognized in clinical practice,^2,3^ with as few as 8% receiving a clinician-documented diagnosis despite meeting guideline diagnostic criteria.^4^ The heterogeneous nature of HFpEF, coupled with imperfect diagnostic tools, contributes to this gap and may delay the initiation of potentially prognostic therapies. ^5^

The American Heart Association has called for the application of artificial intelligence (AI)– driven methods,^6^ particularly unsupervised learning, to better define clinically and biologically meaningful HFpEF subtypes. Its recent Scientific Statement on the Primary Prevention of Heart Failure further highlighted need to shift focus to early-HF stages by leveraging electronic health record (EHR) data to better understand disease progression and thereby enable timely preventive therapy.^7^ Previous studies have derived distinct HFpEF phenotypes using clinical trial datasets. ^8–13^ Critically however, their generalizability is limited by trial-specific inclusion criteria, variable ejection fraction thresholds and reliance on hospitalization-based diagnostic codes. ^8–10,13–15^ These factors introduce selection bias and risk overlooking earlier stages of disease, which may be modifiable.^6^ Furthermore, while prior work has described HFpEF phenotypes at a single point in time, little is known about whether these phenogroups—distinct at presentation—follow different disease trajectories, whether patients remain within assigned phenogroups, or whether some subgroups progress more rapidly than others.^16^ Understanding these patterns is key to identifying high-risk or potentially modifiable disease states.

In this study, we applied an AI-enabled phenotyping framework to a large, real-world EHR dataset. Using natural language processing (NLP), we identified HFpEF cases directly from unstructured clinical text, thereby overcoming limitations of ICD-based diagnosis. We used unsupervised clustering to derive phenogroups at diagnosis. We also utilized longitudinal data to track phenogroup transitions and outcomes. Our goal was to identify novel early-stage subtypes, characterize distinct HFpEF progression trajectories, and evaluate the reproducibility of these findings in an independent external cohort.

## Methods

This retrospective, observational study operated under London South-East Research Ethics Committee approval (18/LO/2048) granted to the King’s Electronic Records Research Interface (KERRI). Individual patient consent was not required. The study complies with the Declaration of Helsinki. The data that support the findings of this study are available from the corresponding author upon reasonable request.

### Derivation cohort

We used an extensively validated NLP pipeline to establish a retrospective database of adult patients with a diagnosis of HFpEF from EHR at the multi-site King’s College Hospital NHS Foundation Trust (KCH) from 2010-2022.^4,17,18^ The original derivation of our cohort has been published in detail previously.^4^ In brief, the diagnosis of HFpEF was based on the latest ESC guidelines.^19^ Patient inclusion followed a structured, stepwise approach, with patients having to fulfil all criteria **(Supplemental Figure S1)**:

1. Clinician-documented heart failure (≥2 mentions of “heart failure” or “HF” in the EHR, identified via NLP).
2. LVEF ≥ 50% within 12 months of the first HF mention. Patients were excluded if they had a recorded LVEF <50% at any time before or after the index HF mention.
3. At least one marker of diastolic dysfunction/raised LV filling pressures: NT-proBNP ≥ 125 pg/ml (≥ 375 pg/ml with AF), LV mass index ≥ 95 g/m^2^ (Female) 115 g/m^2^ (Male), left atrial (LA) volume index > 34 mL/m^2^ (> 40 mL/m^2^ with AF), E/e’ ratio > 9, pulmonary artery systolic pressure (PASP) >35mmHg or tricuspid regurgitation (TR) velocity >2.8m/s.^19^

### Data extraction

MedCAT (Medical Concept Annotation Tool) NLP tools within the CogStack platform were used to extract SNOMED-CT (Systematized Nomenclature of Medicine - Clinical Terms) conceptsP from complete EHR, ^20,21^ including all inpatient documentation, clinic letters, discharge summaries and referrals. In addition to identifying mentions of HF, the NLP pipeline extracted explicitly documented “HFpEF” or “heart failure with preserved ejection fraction”, as well as comorbidities (type 2 diabetes mellitus, hypertension, chronic kidney disease (CKD), atrial fibrillation (AF), myocardial infarction, coronary artery disease (CAD), cerebrovascular accident (CVA; stroke or transient ischemic attack)) and medications (loop diuretics, angiotensin-converting enzyme inhibitors, angiotensin II receptor blockers, beta-blockers and calcium channel blockers). The tool accounted for synonyms e.g. ‘HF’ and applied contextual filters to exclude negations, hypothetical mentions and terms referring to individuals other than the patient. The date of diagnosis was defined as the first mention of heart failure. Previous manual validation of a random representative sample of 100 NLP-extracted ’HFpEF’ mentions demonstrated a 100% true-positive rate for correctly identifying clinician-recorded HFpEF documentation.^18^ Structured data provided demographics, body mass index (BMI), echocardiography reports, natriuretic peptides, hemoglobin, creatinine and HbA1c. For analysis, BNP (B-type natriuretic peptide)/NT-proBNP (N-terminal pro-B-type natriuretic peptide) levels (available in 70% of the cohort) were combined into a single log-transformed z-score, following established methodology.^9^ H_2_FPEF and HFpEF-ABA scores were calculated from baseline data.^22,23^ Mortality data were obtained from EHR death notification letters. As per the stepwise approach, after confirmation of a clinician documented heart failure, the classification of HFpEF was determined independently using structured EHR data according to ESC criteria, irrespective of whether HFpEF was explicitly clinician documented in clinical notes. The final cohort included a subset of patients who met ESC HFpEF criteria but lacked explicit documentation of ’HFpEF’ in their records. By applying objective criteria uniformly across all patient records, this approach mitigates the risk of sampling bias caused by the under-recognition of HFpEF in routine clinical practice. This distinction is critical, as formal clinical recognition of HFpEF is the first step toward appropriate referral, specialist evaluation, and access to guideline-directed therapies. See **Supplemental Methods S1** for more detail.

### Clustering approach and variable selection

Phenogroups were identified using LCA, an unsupervised clustering approach that applies maximum likelihood estimation to classify individuals into mutually exclusive subgroups based on shared characteristics. By identifying latent subgroups through associations between multiple clinical features rather than a single trait, LCA is particularly well suited for HFpEF phenotyping, where a combination of risk factors contributes to disease manifestation. For these reasons this method has been widely applied to HFpEF research.^8,10,14,15^ Alternative methods, including hierarchical clustering and model-based approaches, were tested but generated an excessive number of subgroups, limiting their clinical applicability.

A total of 10 variables were selected for clustering. Variables were selected a priori based on known associations with HFpEF prognosis and clustering approaches in prior studies. 6,7,9,13 Variables included age (<65 years, 65–75 years, 75–85 years, and >85 years), sex, body mass index (BMI) (World Health Organisation classifications: <18.5 kg/m^2^, 18.4-25 kg/m^2^, 25–30 kg/m^2^, >30 kg/m^2^), hypertension, AF, diabetes, CKD, CAD and smoking history and natriuretic peptide quartiles. Missing baseline continuous data (**Supplemental Table S1**) were imputed using the *missForest* package in R. LCA was implemented using the *poLCA* package in R,^24^ with model estimation iterated 20 times across 2–10 potential subgroups. The optimal number of phenogroups was determined using the first Bayesian Information Criterion (BIC) minima (**Supplemental Table S2**). Posterior class membership probabilities were generated for each individual **(Supplemental Table S3)**, representing the likelihood of belonging to each phenogroup given an observed characteristic. Patients were then assigned to the phenogroup with the highest posterior probability, corresponding to maximum-likelihood classification **(Supplemental Table S4)**.

### Longitudinal analysis

To analyze phenogroup transitions over time, we leveraged the temporal data available within our NLP-extracted dataset. Each clinical term was tagged with a timestamp, enabling precise identification of when new diagnoses first appeared in patient records. New diagnoses were therefore defined as comorbidities first documented after the index HF diagnosis date. We implemented a two-step approach to track patient phenogroup transitions. First, a supervised Random Forest (RF) classifier was trained on baseline LCA variables to predict phenogroup membership. The model was optimized using 5-fold cross-validation to minimize out-of-bag error, with variable importance assessed (**Supplemental Figure S2**).

Second, at each annual timepoint from first HF mention, we updated each patient’s clinical profile to include any newly documented comorbidities that appeared in the EHR during that interval. For patients with irregular follow-up schedules, we used the most recent clinical data available within ±2 months of each annual timepoint. The RF model was then applied to these updated profiles to determine phenogroup membership at each timepoint up to 5 years. This approach allowed us to observe how patients transitioned between phenogroups as their clinical characteristics changed over time. Death was treated as a separate competing outcome, allowing us to distinguish between phenotype transitions and mortality outcomes.

Two validation measures were employed: firstly, phenogroup stability was assessed using patients with <10% change in LCA model variables from baseline, and secondly model confidence was evaluated using RF-assigned probabilities, with high confidence defined as >90%.

### External validation

We validated the phenogroups by independently applying the same LCA approach to an independent external cohort from Guy’s and St. Thomas’ NHS Foundation Trust (GSTT), derived using identical inclusion criteria and NLP methods. Phenogroups number was guided by BIC and phenogroup similarity was evaluated by comparing clinical, biochemical, and echocardiographic characteristics.

### Statistical analysis

Baseline continuous variables were reported as mean ± standard deviation, or median with interquartile range (IQR); categorical data were summarized as counts and percentages. Continuous variables were compared with a t-test or Kruskal–Wallis test, categorical variables were compared with a Chi-square test. Survival analysis employed Kaplan-Meier estimates with log-rank tests and Cox proportional hazards models, adjusted for age and sex. The results of the Cox regression analysis are presented as hazard ratios (HRs) with corresponding 95% confidence intervals (CIs). P values <0.05 were designated statistically significant. All analyses were performed using R version 4.4.0.

## Results

We identified 2,223 patients with HFpEF, with only 231 (10.4%) having clinician-documented HFpEF. Prevalent comorbidities included hypertension (67%), CKD (33%), diabetes (26%), CAD (25%), and AF (23%). Median BMI was 29 kg/m² (IQR 25-34) and LVEF was 60.1% (IQR 58.1-62.5). LCA identified four distinct phenogroups (**Figure 1**, **Table 1**).

**Figure 1.**
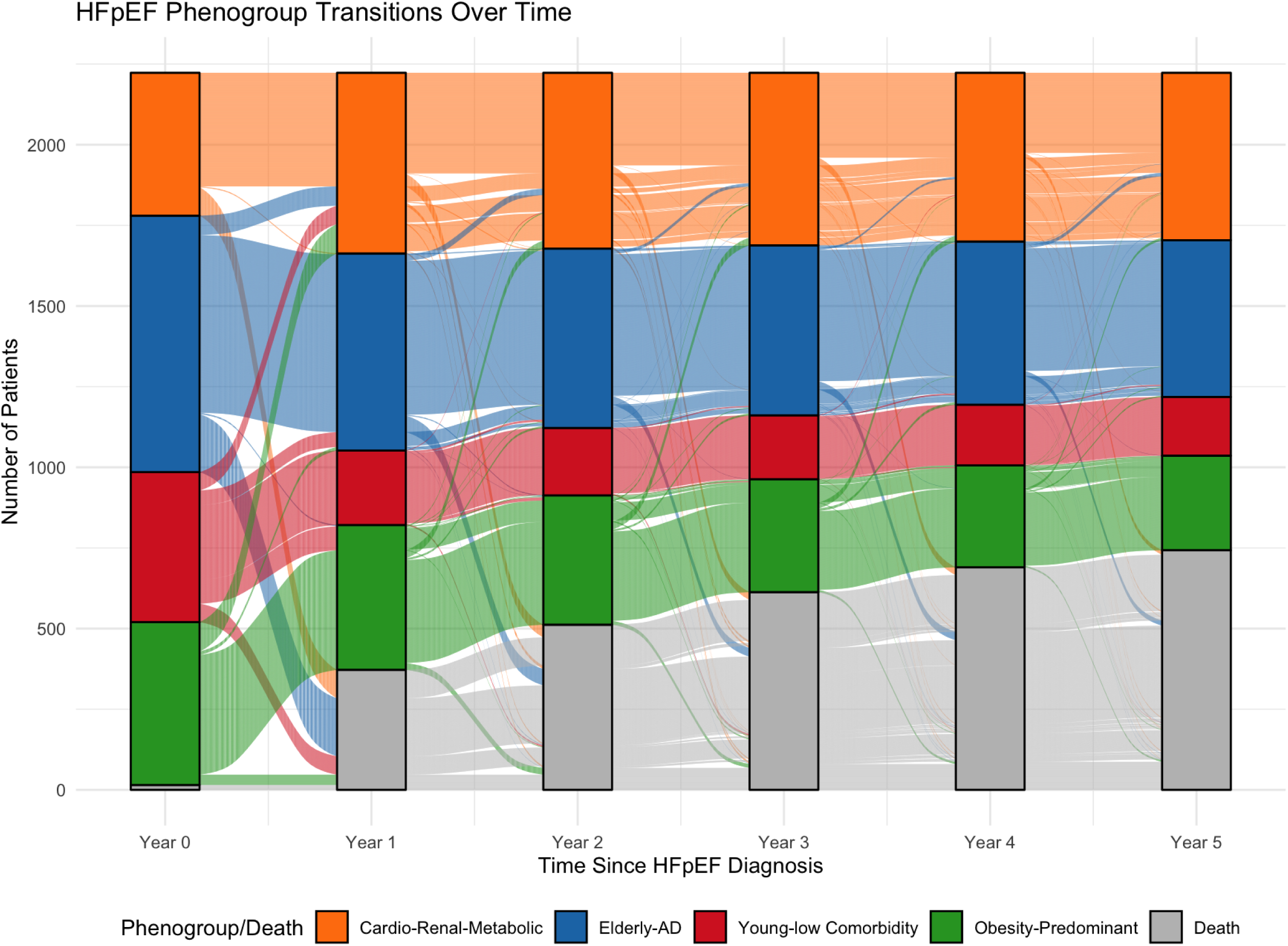
Clinical features of the four phenogroups. Radar plots depicting the relative differences in clinical features across the four phenogroups. Categorical variables are shown as percentages, and continuous variables are displayed as normalized means of Z-standardized values. Abbreviations: AF, atrial fibrillation; CAD, coronary artery disease; CKD, chronic kidney disease; LAVI, left atrial volume index; IVSd, interventricular septum thickness at end-diastole; LVMI, left ventricular mass index; TVR, tricuspid regurgitation maximal velocity; BNP; B-type natriuretic peptide).

**Table 1:** Baseline Demographics, Clinical, and Laboratory Characteristics of HFpEF Phenogroups.

|  | **Elderly-AD,**<br>**32% (n = 703)** | **Cardio-Renal-**<br>**Metabolic,**<br>**23% (n = 503)** | **Obesity**<br>**Predominant,**<br>**24% (n = 530)** | **Young Low**<br>**Comorbidity,**<br>**22% (n= 487)** | **p-value** |
| --- | --- | --- | --- | --- | --- |
| Clinician diagnosis | 78 (11%) | 70 (14%) | 68 (13%) | 15 (3%) | **<0.001** |
| Age | 84 (81, 88) | 74 (65, 81) | 70 (61, 76) | 60 (48, 71) | **<0.001** |
| Female | 482 (69%) | 253 (50%) | 339 (64%) | 257 (53%) | **<0.001** |
| Obesity | 58 (8%) | 321 (62%) | 413 (78%) | 209 (43%) | **<0.001** |
| BMI (kg/m<sup>2</sup>) | 25 (22, 28) | 31 (28, 36) | 33 (30, 38) | 29 (25, 34) | **<0.001** |
| Ethnicity |  |  |  |  |  |
| White | 484 (70%) | 248 (48%) | 257 (49%) | 272 (55%) | **<0.001** |
| Black | 101 (15%) | 168 (33%) | 181 (34%) | 128 (26%) | **<0.001** |
| Asian | 30 (4.4%) | 46 (8.9%) | 36 (6.9%) | 22 (4.4%) | **0.003** |
| Hypertension | 487 (69%) | 480 (95%) | 498 (94%) | 14 (2.9%) | **<0.001** |
| Atrial Fibrillation | 264 (38%) | 138 (27%) | 95 (18%) | 17 (3%) | **<0.001** |
| CVA | 174 (25%) | 181 (35%) | 102 (19%) | 36 (7.3%) | **<0.001** |
| Chronic Kidney<br>Disease | 190 (27%) | 498 (99%) | 27 (5%) | 48 (10%) | **<0.001** |
| Diabetes Mellitus | 70 (10%) | 342 (68%) | 176 (33%) | 11 (2%) | **<0.001** |
| Pulmonary hypertension | 189 (27%) | 168 (33%) | 144 (27%) | 131 (27%) | **0.048** |
| Coronary artery disease | 156 (22%) | 218 (43%) | 172 (32%) | 18 (3.7%) | **<0.001** |
| Smoking history | 453 (66%) | 299 (58%) | 297 (57%) | 333 (67%) | **<0.001** |
| Anaemia | 354 (50%) | 305 (61%) | 190 (36%) | 252 (52%) | **<0.001** |
| Haemoglobin | 121 (108, 133) | 117 (101, 132) | 130 (115, 140) | 122 (101, 136) | **<0.001** |
| Creatinine | 87 (70, 114) | 125 (90, 203) | 79 (64, 96) | 75 (59, 97) | **<0.001** |
| Hypotensive agent | 428 (61%) | 448 (89%) | 391 (74%) | 100 (21%) | **<0.001** |
| Loop diuretics | 316 (52%) | 339 (75%) | 219 (56%) | 84 (29%) | **<0.001** |
| Beta blockers | 267 (38%) | 303 (60%) | 206 (39%) | 66 (14%) | **<0.001** |
| CCBs | 214 (30%) | 336 (67%) | 252 (48%) | 25 (5%) | **<0.001** |
| ACE inhibitors | 305 (43%) | 377 (75%) | 307 (58%) | 46 (9%) | **<0.001** |
| NT-proBNP | 3,562 (2,128, 5,349) | 3,824 (1,506, 7,298) | 987 (404, 1,853) | 1,775 (971, 3,134) | **<0.001** |
| H<sub>2</sub>FPEF score | 6 (3, 6) | 5 (3, 6) | 4 (3, 6) | 3 (2, 4) | **<0.001** |
| HFpEF-ABA score | 81 (67, 94) | 81 (64, 92) | 75 (59, 89) | 51 (29, 72) | **<0.001** |

Median (IQR) or n (%)
Abbreviations: BMI, Body Mass Index; CVA, Cerebrovascular accident; NT-proBNP, N- terminal B-type natriuretic peptide.

### Phenogroup characteristics

Phenogroup 1 (Elderly - Atrial Dysfunction (AD)) was the largest (n = 703) and oldest (median 84 years, IQR 81-89), predominantly female (69%) and White (70%), with highest prevalence of AF (38%), largest LA volumes (42 ml/m^2^, IQR 33-46), pulmonary artery pressures (31 mmHg IQR (25-41) and highest H_2_FPEF and HFpEF-ABA scores (**Table 1 and 2**). Phenogroup 2 (Cardio-Renal-Metabolic) had the greatest comorbidity burden, with a high prevalence of CKD (99%), hypertension (95%) and diabetes (68%). The group also had the highest NT-proBNP (3,824 pg/ml), LV mass (106 g/m^2^ IQR 93-121) and greatest diastolic dysfunction. Additionally, it had the most clinician-documented HFpEF (14%). Phenogroup 3 (Obesity-Predominant) had the highest obesity prevalence (78%) and substantial hypertension (95%), but lower rates of diabetes (33%) and CKD (5%) than the Cardio-Renal-Metabolic phenogroup, as well as significantly lower NTproBNP concentrations (987 pg/ml IQR (404-1,853)) but a similar number of clinician-documented cases (13%). Finally, phenogroup 4 (Young-Low Comorbidity) was the youngest (median 60 years, IQR 48-71) with minimal comorbidities (hypertension 3%, AF 3%, diabetes 2%) and mildest echocardiographic abnormalities. NTproBNP concentrations were raised at 1,775 pg/ml IQR (971 - 3,134). This group had the fewest clinician-documented HFpEF diagnoses (3%).

**Table 2:**
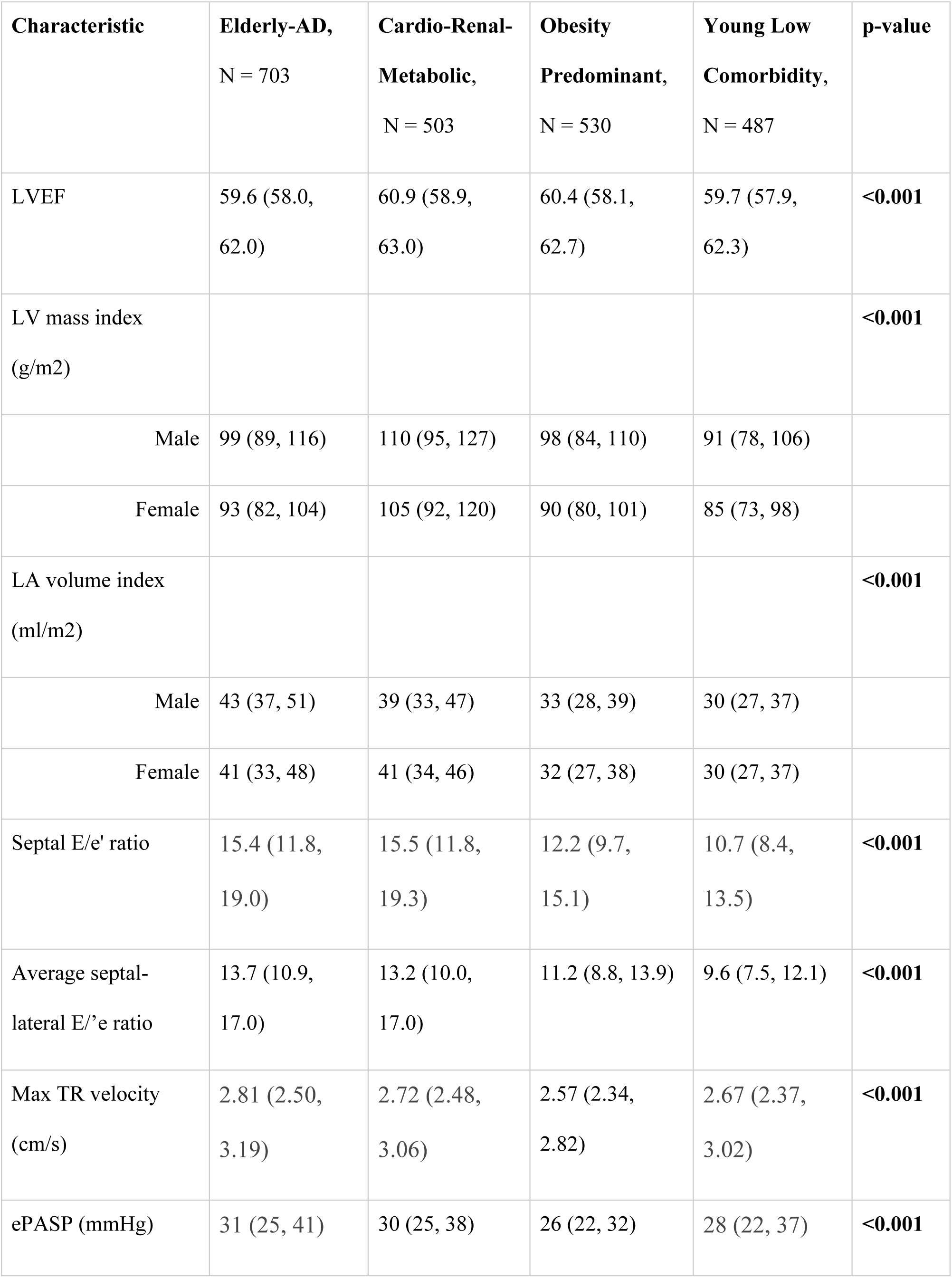
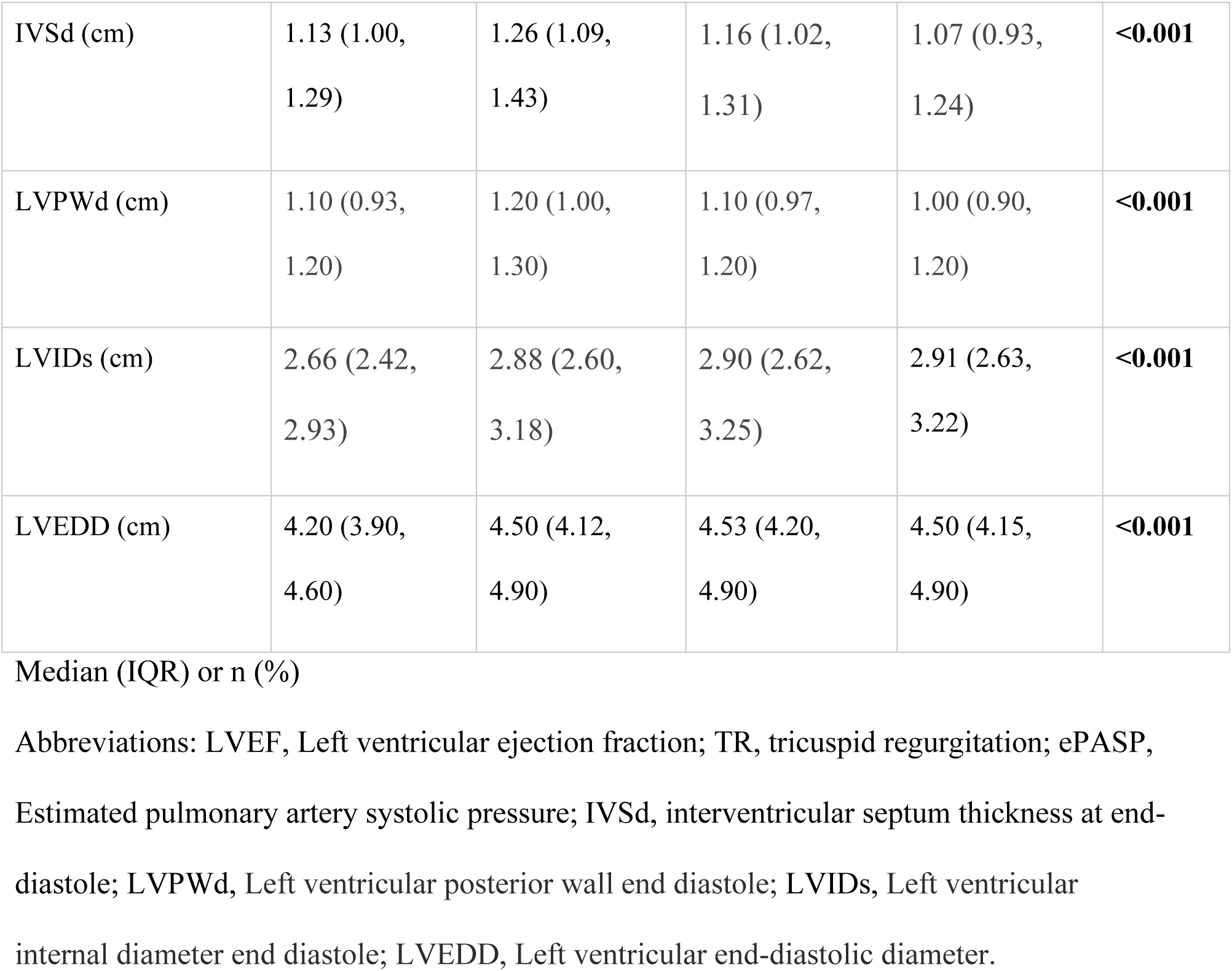
Echocardiographic Parameters Across HFpEF Phenogroups.

### Phenogroup trajectories

Over a median follow-up of 4.0 years (IQR 2.22-6.10), we tracked yearly phenogroup reassignments. Our RF model demonstrated high stability (92% consistency in patients with stable clinical characteristics) and confidence (mean score 0.90±0.13). Phenogroup transitions (**Figure 2**) revealed several patterns: The Cardio-Renal-Metabolic group experienced the highest stability (<1% transitions), while the Elderly-AD group demonstrated modest movement to Cardio-Renal-Metabolic (7.6% at year 1). The Obesity-Predominant group demonstrated moderate transition rates, primarily shifting toward Cardio-Renal-Metabolic after two years (peaking at 28.3% by year 5). The Young-Low Comorbidity group exhibited greatest movement, transitioning to Obesity-Predominant (15.9% at year 1) and Cardio-Renal-Metabolic (12.5% by year 3). Patients moving from the Young-Low Comorbidity phenogroup were older at baseline (66 vs. 54 years, p <0.001) and had a higher LV mass (95g/m^2^ (IQR 81 - 110g/m^2^) vs. 82g/m^2^ (IQR 70 - 95/m^2^), p <0.001) and septal E/e’ ratio (13.0 (IQR 10.0 - 16.1) vs 10.0 (7.3 - 12.7), p <0.001). The greatest phenotypic shifts occurred within the first two years, after which transitions stabilized.

**Figure 2.**
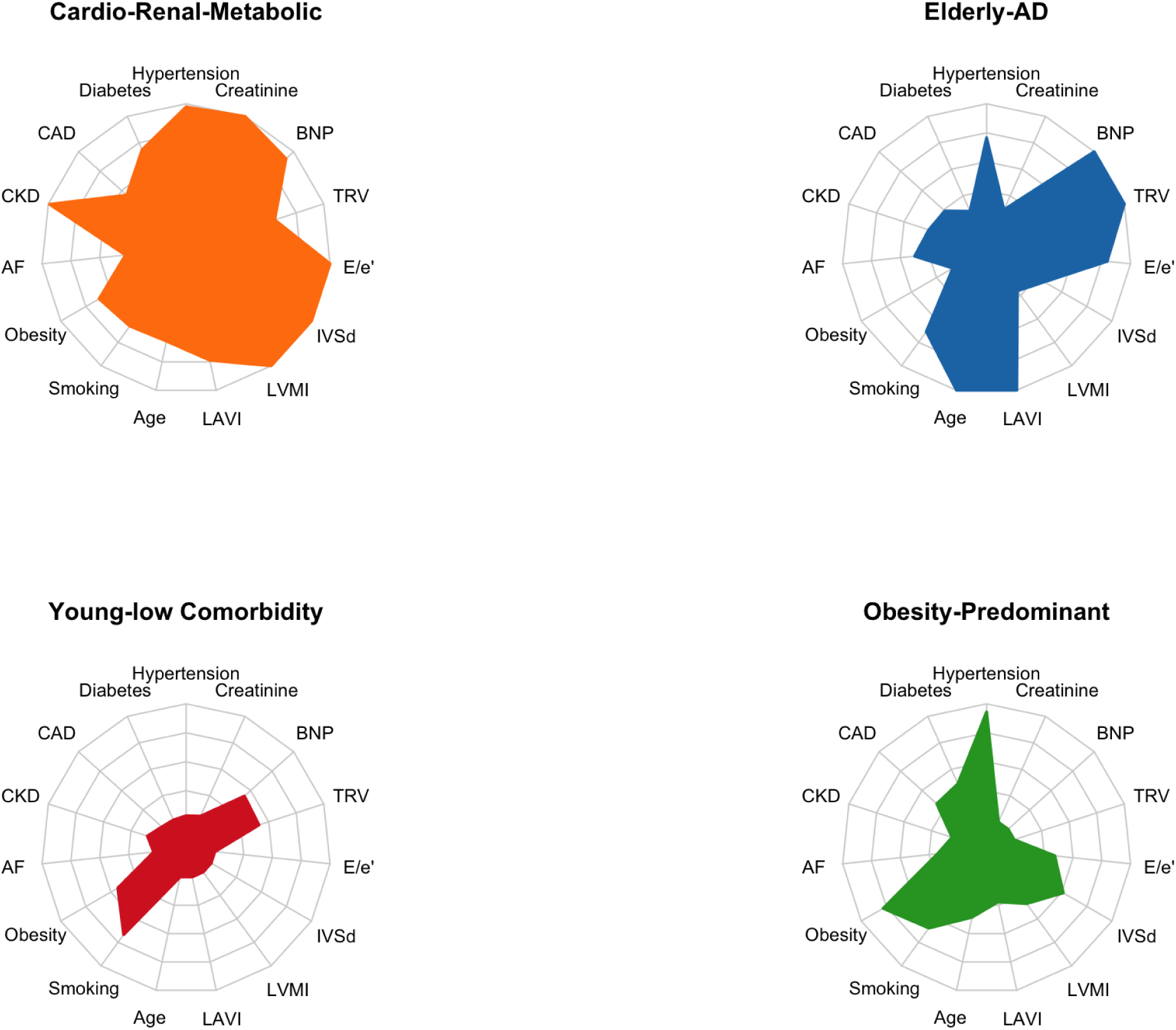
HFpEF phenogroup trajectories. Alluvial diagram depicting transitions between HFpEF phenogroups (Cardio-Renal-Metabolic, Elderly-AD, Young-Low Comorbidity, and Obesity-Predominant) across 5 years following initial diagnosis. Phenogroup membership was reclassified annually using a supervised Random Forest model trained on baseline LCA variables and applied to updated clinical characteristics. For each timepoint (1, 2, 3, 4, and 5 years), the most recent data within ±2 months were used. The width of each flow represents the proportion of patients transitioning between phenogroups. Death is modelled as a competing event, with transitions reflecting patients who remained alive.

### Survival

Overall, 5-year survival was 64% (95% CI 62-66%) (**Figure 3A, Supplemental Table S5**). After age-and-sex adjustment, the Elderly-AD (HR 1.30, 95% CI 1.02-1.67, p=0.03) and Cardio-Renal-Metabolic phenogroups (HR 1.49, 95% CI 1.18-1.87, p<0.001) demonstrated significantly higher mortality risk compared to the Young-Low Comorbidity reference group (**Figure 3B**). No significant mortality difference was observed between Young-Low Comorbidity and Obesity-Predominant phenogroups.

**Figure 3.**
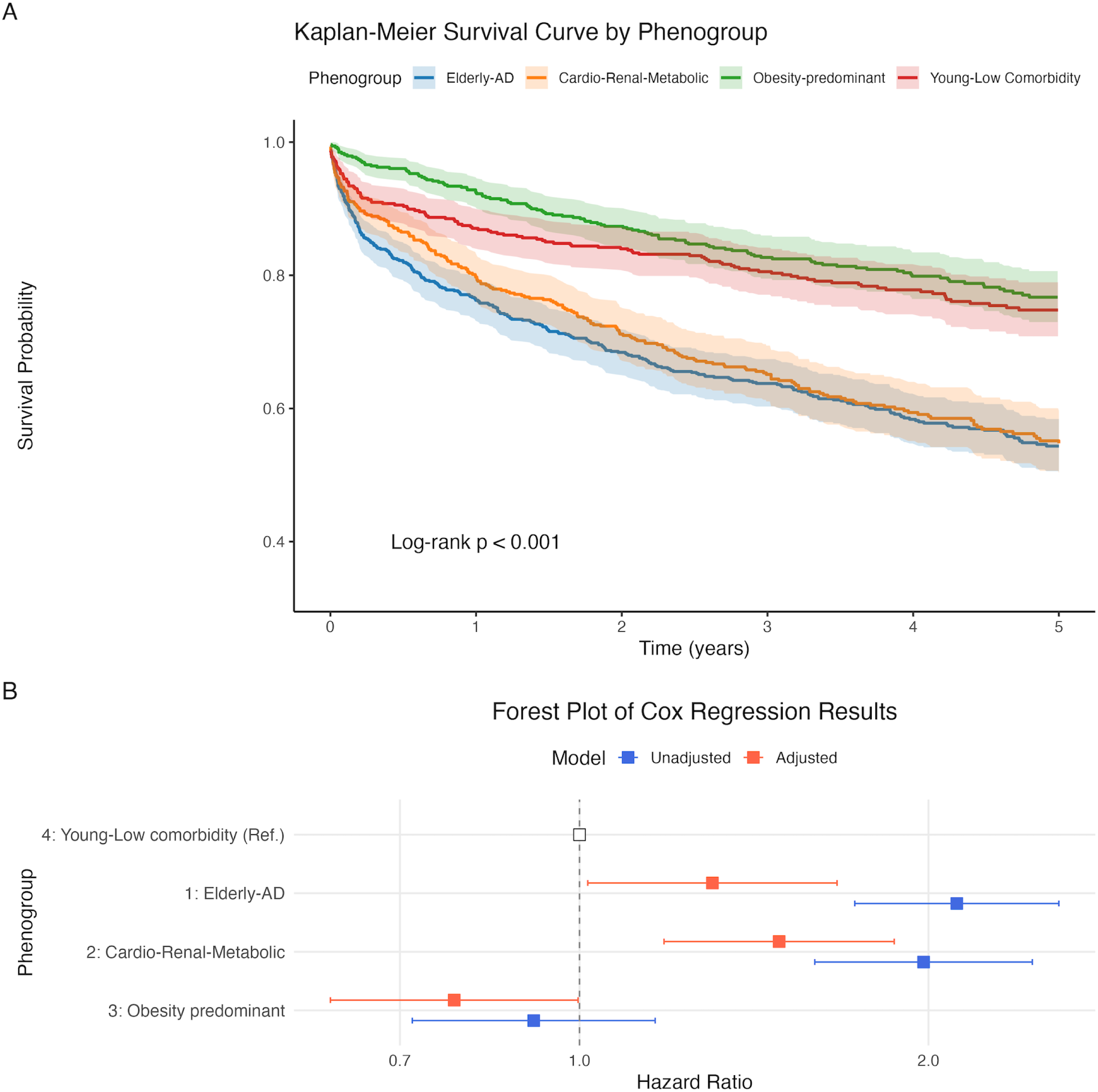
Kaplan-Meier Survival Curves and Forest Plot of Adjusted Hazard Ratios for HFpEF Phenogroups. *(A)* Kaplan-Meier survival curves illustrating five-year survival estimates stratified by HFpEF phenogroup. Log-rank tests were used to assess significant differences in survival across phenogroups. *(B)* Forest plot displaying adjusted and unadjusted hazard ratios (HR) for all-cause mortality across HFpEF phenogroups, with 95% confidence intervals (CI). The Young-Low Comorbidity phenogroup served as the reference group. Adjusted for age and sex.

### External validation

External validation in the GSTT cohort (n=3,349) confirmed our findings despite higher baseline comorbidity prevalence (diabetes 36% vs. 26%, AF 42% vs. 23%, CKD 43% vs. 33%). LCA identified four clusters with similar clinical characteristics and mortality trends. (**Supplemental Table S6, Supplemental Figure S3**). The Elderly-AD phenogroup showed the highest AF prevalence in both cohorts (KCH 38%, GSTT 92%) and significantly elevated mortality risk (unadjusted HR 2.07, 95% CI 1.76-2.44, p<0.001). The Cardio-Renal-Metabolic (HR 1.46, 95% CI 1.27-1.69, p<0.001) and Obesity-Predominant phenogroups (HR 1.20, 95% CI 1.02-1.48, p=0.03) also showed increased mortality compared to the Young-Low Comorbidity reference group.

## Discussion

The main findings of our study are as follows: 1) We demonstrated that an NLP-enabled interrogation of EHRs, combined with unsupervised clustering, successfully identified four distinct phenogroups within a real-world HFpEF cohort; 2) Unlike previous cross-sectional work, our longitudinal approach revealed dynamic phenogroup transitions, distinguishing both stable end-stage phenotypes and earlier-stage, potentially modifiable disease; 3) We identified a prognostically relevant, under-recognized early-stage HFpEF, representing a key target for earlier detection and intervention.

### Phenogroup characterization and transitions

Our results extend previous phenotyping work, by integrating longitudinal data to track phenogroup transitions over time, thereby addressing a key limitation of existing HFpEF literature. Prior studies have primarily relied on clinical trial populations, ^8–10,13–15^ with restrictive inclusion criteria and variable diagnostic thresholds, limiting generalizability to broader clinical practice. In contrast, our application of latent class analysis to a real-world, NLP-identified cohort meeting ESC HFpEF criteria enabled the capture of phenotypes across a wider clinical spectrum, including patients without a formal HFpEF label. This not only reduces selection bias but reflects the under-recognition of HFpEF in routine care, ^2^ with implications for future HFpEF trial design. These insights align with recent AHA scientific statements calling for more refined approaches to HFpEF classification,^6^ particularly through use of EHR data to better understand disease onset and trajectory. Such approaches are especially relevant given the absence of a unifying diagnostic marker in HFpEF, the high burden of comorbidities that may obscure its detection. ^2^

The Young–Low Comorbidity group represents an under-recognized HFpEF phenotype in clinical practice with important implications. Despite the documentation of heart failure, meeting ESC diagnostic criteria and having elevated NT-proBNP, patients in this group were infrequently labelled as HFpEF within their clinical notes. While previous studies have identified younger, lower-risk subgroups ^10,12,13,15^ our study extends these findings by demonstrating that such patients frequently transition to higher-risk phenotypes— most notably to the Cardio-Renal-Metabolic group (12.5% by year 3) (**Figure 2**). These patterns suggest the presence of a distinct early-stage HFpEF, potentially influenced by contributors such as central adiposity or inflammation, which may not be adequately captured by BMI alone. ^25^ Importantly, patients within this early phenogroup still experienced substantial five-year mortality (26%), underscoring that adverse outcomes can arise even in phenotypically mild presentations. This highlights a critical window for intervention, where therapies targeting metabolic or inflammatory pathways may delay or prevent progression to more advanced disease.

The Obesity-Predominant phenogroup was characterized by younger age, the highest BMI, preserved renal function, and moderate LV hypertrophy. Despite lower natriuretic peptide levels, structural remodeling was evident, consistent with previously described obesity-related HFpEF. ^26^ Although initially stable, this group showed delayed but substantial progression to the Cardio-Renal-Metabolic phenotype by year 5 (28.3%), consistent with the accumulation of metabolic risk factors over time.

The Cardio-Renal-Metabolic phenotype is a well-established high-risk HFpEF phenotype marked by a high prevalence of diabetes and CKD, LV hypertrophy, elevated filling pressures, and natriuretic peptides. ^9,11–13^ This group had the worst five-year mortality and demonstrated phenotypic stability (<1% annual transitions), consistent with an end-stage disease state. The phenotype aligns closely with the recently proposed cardiovascular-kidney-metabolic (CKM) syndrome framework, highlighting the intertwined pathophysiology and risk conferred by metabolic, renal, and cardiovascular dysfunction.^27^ Recent evidence suggests that the combination of CKD and diabetes, both independently associated with adverse cardiac structure and prognosis in HFpEF ^28,29^, may act synergistically to drive the excess mortality observed in this phenotype. ^30^ This mechanistic interplay may partly explain the significant divergence in outcomes compared to the Obesity-Predominant phenotype, which lacks the same degree of CKM burden.

The Elderly–Atrial Dysfunction group exhibited pronounced diastolic dysfunction, marked LA enlargement, high rates of AF, and elevated pulmonary pressures. These features are characteristic of atrial myopathy phenotypes seen in older HFpEF patients.^9,11,13,31^ Atrial fibrillation and HFpEF often coexist, and many symptomatic AF patients, even without an HF diagnosis, demonstrate elevated filling pressures and early HFpEF physiology.^3,32^ In our cohort, this group had the most severe atrial remodeling and high-HFpEF scores, with minimal transitions and high mortality, suggesting late-stage phenotype. Together, these findings reinforce HFpEF as a progressive syndrome. The Young–Low Comorbidity group represents a potential early phase, while the Elderly–Atrial Dysfunction and Cardio-Renal-Metabolic phenogroups reflect end-stage disease. Prior studies have described Stage D HFpEF subtypes marked by severe atrial dysfunction, metabolic disease, and multi-organ involvement, features captured in our advanced high mortality phenogroups. ^33^ Identifying and targeting early-stage phenotypes may be key to preventing progression to these poor-prognosis states, since disease stage may in part determine treatment response.^34^

### Implications for early detection and intervention

Despite all patients in our cohort meeting standardized objective HFpEF criteria, many remained clinically unrecognized, highlighting persistent challenges of diagnosing HFpEF. Tools such as HFpEF-ABA,^23^ can aid detection, but our study highlights its limited discrimination in younger patients with a lower comorbidity burden. The widespread adoption of EHRs presents an opportunity to implement AI-based multi-modality diagnostic tools into routine clinical workflows that reduce reliance on clinician suspicion to facilitate earlier HFpEF identification. Despite the central role of echocardiography, it is underutilized in community care, and key HFpEF diagnostic parameters are inconsistently assessed. ^35^ Imaging-integrated EHR diagnostic tools may therefore offer a more accurate and scalable approach, particularly in high-risk populations such as those with obesity or AF.^2,3^ Shifting the diagnosis earlier provides the opportunity for establishing Sodium-glucose co-transporter-2 (SGLT2) inhibitors or Glucagon-like peptide-1 (GLP-1) agonists which have shown benefit in HFpEF and may be more impactful if deployed earlier in the disease course.^36,37^

This study has several key strengths. First, unlike clinician-diagnosed cohorts, our validated NLP approach extracts heart failure diagnoses directly from unstructured clinical text, capturing under-recognized and early-stage phenotypes missed by structured coding systems. By leveraging NLP in this way, we extend its previously demonstrated utility for echocardiography data extraction and outcome adjudication,^38,39^ to the context of HFpEF phenotyping in a large, real-world dataset. Second, the longitudinal design represents an advance over previous cross-sectional studies. Finally, despite differences in baseline characteristics, the reproducibility of phenogroup features in an independent validation cohort and their consistency with prior studies underscore the robustness of our findings.

### Limitations

We acknowledge several limitations. First, this was a retrospective study and while findings were externally validated, further prospective validation across diverse populations from different geographies is warranted. Second, reliance on EHR-based NLP extraction depends on the completeness and accuracy of routine clinical documentation. As such there is potential for under-recording at baseline, which may influence the observed phenogroups and transitions. Additionally cause-specific mortality data was not available. Finally, clustering was based on comorbidities and therefore incorporating additional biomarkers or clinical data may have provided different phenogroups.

## Conclusion

NLP-enabled phenotyping of real-world EHR data identified and externally validated four reproducible HFpEF phenogroups with distinct clinical characteristics and trajectories.

Among these, we uncovered an underrecognized, younger early-stage phenogroup, characterized by minimal comorbidities yet substantial mortality. These findings underscore the diagnostic challenges of early HFpEF and highlight the potential of EHR-based, AI-driven strategies to improve earlier identification and faciliate targeted, guideline-directed therapy.

## Data Availability

The data that support the findings of this study are available from the corresponding author upon reasonable request.

## Abbreviations

NLP: Natural language processing
LCA: Latent class analysis
EHR: Electronic health records

## Acknowledgments

None

